# High-resolution Orbitofrontal Cortex Morphometry and Cannabis Use Disorder Severity in High-risk Emerging Adults: A Preliminary Study

**DOI:** 10.64898/2026.05.26.26354113

**Authors:** Tegan L. Hargreaves, Carly McIntyre-Wood, Mahmoud Elsayed, Emily Vandehei, Kyla L. Belisario, Laura Lee, Ashley Blakely, Jillian Halladay, Michael Amlung, Lawrence H. Sweet, James MacKillop

## Abstract

**Background:** Cannabis use is highly prevalent among emerging adults (18-25 years), a developmental period marked by ongoing neurodevelopment and heightened risk for cannabis use disorder (CUD). Structural alterations in the orbitofrontal cortex (OFC) and medial prefrontal/anterior cingulate cortex (mPFC/ACC) have been linked to cannabis use, though findings remain inconsistent in directionality. To address this, we examined cortical thickness and surface area of the OFC and mPFC/ACC subregions using the high-resolution Glasser atlas, allowing for more granular characterization of associations with CUD severity.

**Method:** One hundred eleven emerging adults (41% male, aged=20.6±1.1 years) reporting significant alcohol and/or cannabis use completed clinical assessments and structural MRI. The OFC and mPFC/ACC were segmented into seven and six subregions per hemisphere, respectively. Multiple linear regressions tested associations between cortical thickness or surface area and DSM-5 CUD symptom count, controlling for alcohol use and intracranial volume. Subregions surviving false discovery rate correction were examined in relation to depression, trauma-related symptoms, impulsivity, and cannabis use motives.

**Results:** Greater CUD severity was associated with lower cortical surface area and greater cortical thickness in OFC and mPFC/ACC subregions. Lower OFC surface area was correlated with coping- and enhancement-related cannabis use motives. Lower mPFC/ACC surface area and greater thickness were associated with more severe depression, trauma-related symptoms, and impulsivity.

**Conclusion:** In high-risk emerging adults, greater CUD symptom burden is associated with lower surface area and greater thickness in OFC and mPFC/ACC subregions. Using the high-resolution Glasser atlas, these findings provide a more precise characterization of structural correlates of CUD and highlight potential neurobiological markers linked to affective and motivational processes underlying cannabis use.

## Introduction

Cannabis is one of the most commonly used substances worldwide, with particularly high prevalence among emerging adults aged 18-25 years (Health Canada, 2023). In Canada, use within this population has steadily increased over time, with substantial increases following the legalization of non-medicinal cannabis use in 2018 and the COVID-19 pandemic (Hall et al., 2023; Doggett et al., 2025; McDonald et al., 2025). Due to its high prevalence and the increased risk of cannabis use disorder (CUD) development in emerging adults, understanding cannabis use at this developmental stage is of utmost importance and represents a major public health concern (Qadeer et al., 2019; Leung et al., 2020; Halladay et al., 2025). Elevated cannabis use and, more critically, CUD in emerging adults have been associated with adverse outcomes such as depression, anxiety, and psychosis, as well as reduced performance in domains such as attention, coordination, decision-making, impulsivity, and memory (Hall et al., 2020; González-Roz et al., 2025; Halladay et al., 2025). This may be driven, in part, by neurodevelopmental changes occurring in emerging adulthood that may be disrupted by significant cannabis use, making this age group particularly vulnerable to poor outcomes (Hall et al., 2020). Therefore, a more comprehensive understanding of CUD during emerging adulthood requires consideration of the impact on brain structure, as well as the factors motivating cannabis use.

A growing body of work has examined associated differences in brain structure during emerging adulthood. Relative to cannabis-free controls, emerging adults who use cannabis tend to demonstrate lower gray matter volume, particularly within frontal regions such as the prefrontal cortex (PFC) and orbitofrontal cortex (OFC), as well as in subcortical structures including the thalamus, hippocampus, and amygdala (Lichenstein et al., 2022; Colyer-Patel et al., 2024; Nosko et al., 2025). Cannabis use has also been associated with alterations in cortical morphometry, including both cortical thickness and surface area (Lichenstein et al., 2022; Colyer-Patel et al., 2024; Nosko et al., 2025). Several studies have reported associations between regular cannabis use or CUD and altered gray matter volume, cortical thickness, or surface area within frontal regions, most consistently implicating the OFC and medial PFC (mPFC)/anterior cingulate cortex (ACC). Reported effects include thinner, smaller surface area, and reduced volume in the OFC, thinner mPFC, and smaller ACC volume (Shollenbarger et al., 2015; Chye et al., 2017; Lorenzetti et al., 2019; Maple et al., 2019; Albaugh et al., 2021; Harper, Wilson, et al., 2021; Li et al., 2025); however, there are some studies that find opposing directionality of reported effects. For example, Jacobus et al. reported greater cortical thickness in the OFC was associated with more severe cannabis use, and Wade et al. found that larger OFC volume predicted CUD status (Jacobus et al., 2015; Wade et al., 2019). Overall, the extant literature demonstrates that cannabis use and CUD status are associated with structural variations, specifically within frontal brain regions that support reward processing and cognition, but the directionality of the effects remains unclear. These discrepancies may reflect the size of the structures as well as their roles in multiple different cognitive domains, warranting further investigation with greater specificity within regions.

Therefore, in the present study, we investigated structural gray matter neural correlates of cannabis use in emerging adults from within the community. For the first time, we used the higher resolution Glasser atlas (Glasser et al., 2016) to parcellate the brain, rather than the traditional Desikan or Destrieux atlases. The Glasser atlas divides the brain into 180 regions per hemisphere, offering a more precise assessment of larger neuroanatomical regions and better insight into diverging existing results. Based on the prevailing literature implicating the OFC (Chye et al., 2017; Lorenzetti et al., 2019; Albaugh et al., 2021; Harper, Wilson, et al., 2021; Li et al., 2025), we were primarily interested in the associations between cortical thickness and surface area of OFC subregions in relation to CUD severity. A secondary priority was examining associations between cortical thickness and surface of mPFC/ACC subregions in relation to CUD severity (Shollenbarger et al., 2015; Maple et al., 2019). Consistent with previous literature, we hypothesized that participants endorsing more severe CUD (i.e., higher number of symptoms) would demonstrate thinner and lesser surface area across larger neuroanatomical regions. Last, among brain regions that were significantly associated with CUD, we examined their relationship with indicators that might inform their functional mechanistic relations with CUD, such as internalizing disorder symptoms (i.e., depression, anxiety, trauma), motives for using cannabis, and impulsivity. We further hypothesized that these relationships would be coupled with more severe internalizing features and higher levels of impulsivity (Hall et al., 2020; González-Roz et al., 2025; Halladay et al., 2025).

## Method

### Participants

Adults aged 19-22 years were recruited from the general community in Hamilton, Ontario, Canada as part of a larger longitudinal study investigating high-risk substance use in emerging adults, focusing on the two most commonly used substances, alcohol and cannabis. Therefore, participants were required to meet the following inclusion criteria: (1) Alcohol Use Disorder Identification Test (AUDIT) score >8 and 3+ heavy drinking days (≥ 5/4 standard drinks for males/females; in the past month) and/or Cannabis Use Disorder Identification Test (CUDIT) score >8 and using cannabis at least twice per week (in the past month); (2) right-handed; and (3) fluent English speaker. Participants were permitted to endorse significant consumption of both alcohol and cannabis. Exclusion criteria included: (1) weekly or greater use of recreational substances other than alcohol, cannabis, or tobacco; (2) history of severe mental health disorder (e.g., schizophrenia, bipolar disorder, etc.); (3) significant history of brain injury (e.g., repeated concussions, traumatic brain injury, stroke, etc.) or other neurological disorder (e.g., multiple sclerosis, epilepsy, etc.); and (4) MRI research contraindications (e.g., pregnancy, metal implants, claustrophobia). All participants provided written informed consent, and the study procedures were approved by the Hamilton Integrated Research Ethics Board (#13763).

A total of 114 participants completed the baseline and MRI sessions. Three participants were excluded due to incidental findings (n=2, *mega cisterna magna*; n=1, *cavum septum pellucidum et vergae*), resulting in 111 participants in the final sample (Table 1).

**Table 1.** Participant Characteristics (n = 111)

| Indicator | Mean (SD) |
| --- | --- |
| Age | 20.57 (1.11) |
| Sex (male; n (%)) | 45 (41) |
| Race (European White, n (%)) | 74 (67) |
| Years of Education | 14.06 (1.87) |
| Income (median) | \$75,000-90,000 |
| Smoker (n (%))* | 41 (37) |
| AUDIT Score | 8.64 (6.22) |
| CUDIT Score | 14.64 (7.69) |
| CUD Symptom Count** | 3.85 (3.07) |
| *CUD Severity (n (%))* |  |
| None (0-1 symptoms) | 32 (28) |
| Mild (2-3 symptoms) | 25 (23) |
| Moderate (4-5 symptoms) | 18 (16) |
| Severe (6+ symptoms) | 36 (32) |

Notes: Based on \* self-report and confirmed with FTND score > 1; \*\* based on DART.

Of the 111 included individuals, 32% (n=35) met inclusion criteria for both alcohol and cannabis use, while 18% (n=20) and 51% (n=56) participants endorsed only alcohol or cannabis use, respectively. All demographic and clinical characteristics are fully described in Table 1. Overall, participants reported high levels of cannabis use, with 72% of individuals meeting criteria for at least a mild CUD (average symptom count=3.85±3.07).

### Procedures

As part of a larger longitudinal study, interested individuals were initially assessed for eligibility via online screening through REDCap (Harris et al., 2009). All eligible participants completed two in-person visits, one that included clinical and out-of-scanner assessments and the second comprising an MRI.

#### Demographic and Clinical Characteristics

At the first visit, participants were asked to provide demographic information including their age, sex assigned at birth, gender identity, financial information (i.e., household income and socioeconomic status), ethnicity, years of education, and employment status.

Participants were administered assessments of substance use including alcohol and cannabis, which included the AUDIT (Babor & Robaina, 2016), CUDIT (Bonn-Miller et al., 2016), and the Diagnostic Assessment Research Tool (DART), a structured, clinician-rated assessment based on DSM-5 criteria of substance use disorders (Schneider et al., 2022).

To better understand associated features of cannabis use, participants were provided assessments to measure depression (9-Item Patient Health Questionnaire [PHQ-9], (Kroenke et al., 2001)), anxiety (7-Item Generalized Anxiety Disorder assessment [GAD-7], (Spitzer et al., 2006)), trauma (Posttraumatic Stress Disorder Checklist for DSM-5 [PCL-5], (Blevins et al., 2015)), impulsivity (Urgency-Premeditation-Perseverance-Sensation-Seeking-Positive Scale [UPPS], (Cyders et al., 2014)), and motives for cannabis use (Marijuana Motives Measure [MMM], (Simons et al., 1998)).

#### MRI Data Acquisition

Participants were scanned with a 3-Tesla whole-body GE Discovery 750 MRI scanner (General Electric, Milwaukee, WI, USA). For the current study, T1-weighted whole-brain volumes were acquired using a BRAVO sequence with the following parameters: repetition time = 7.4 ms, echo time = 3.06 ms, inversion time = 450 ms, 12° flip angle, and 188 contiguous 1 mm sagittal slices, resulting in 1 mm isometric voxels. As part of the entire study protocol, resting-state and task-based runs were acquired, as well as an MPRAGE T1 structural acquisition, resulting in a total scan time of roughly 45 minutes.

### Data Analysis

T1-weighted structural images were segmented and parcellated using the standard recon-all pipeline in FreeSurfer v8.0 (https://surfer.nmr.mgh.harvard.edu/). Full details outlining this pipeline can be found elsewhere (https://surfer.nmr.mgh.harvard.edu/fswiki/recon-all). All segmentations were inspected by trained raters (TLH, CMW, ME).

Following cortical reconstruction and volumetric segmentation, each participant’s cortical surface was parcellated according to the Human Connectome Project’s multimodal parcellation (i.e., HCP-MMP1.0) (Glasser et al., 2016). Parcellation files were then used to extract vertex-wise morphometric measures of cortical thickness and surface area for each lateralized region (i.e., 180 per hemisphere and measure). Based on previous literature, we focused on the OFC as the primary analysis (Harper, Malone, et al., 2021; Lichenstein et al., 2022; Li et al., 2025). The Glasser atlas segments the frontal poles and OFC into 11 subregions per hemisphere based on structural and functional variations, also corresponding with Brodmann areas (BA) (Glasser et al., 2016). To avoid inflated type I error, we selected only OFC regions of interest—rather than the frontal poles—and assessed both hemispheres (Figure 1A): the OFC, posterior OFC (pOFC), BA 11 (lateral portion of area 11 [11l]), BA 13 (lateral portion of area 13 [13l]), and BA 47 (47s, 47m, anterior portion of 47r). As a secondary region of interest, we selected the mPFC/ACC, which included the BA 8 (area 8BM), BA 9 (middle portion of area 9), and BA 32 (areas a32 prime [a32pr], p32 prime [p32pr], d32, p32) (Figure 1B). Across all analyses, we were interested in the cortical thickness and surface area of the a priori regions, rather than cortical volume, as they provide more specific information about cortical morphometry, and thickness is largely collinear with volume (Durazzo et al., 2011; Hargreaves et al., 2025).

**Figure 1.**
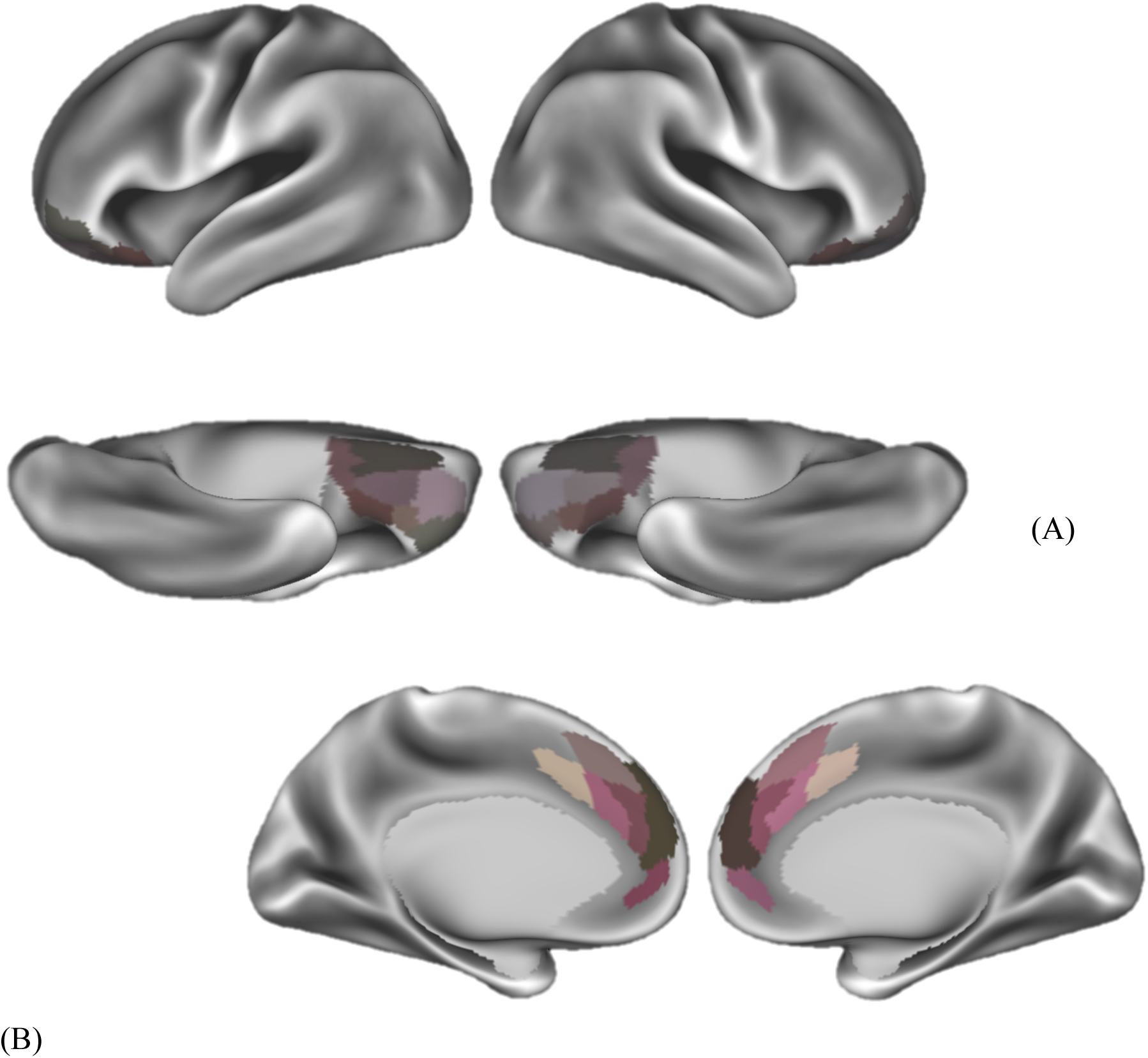
*A priori* regions of interest, per the Glasser atlas. (A) Lateral and ventral views of the orbitofrontal cortex and (B) medial view of the medial prefrontal cortex/anterior cingulate cortex subregions of interest.

### Statistical Analysis

To best understand the relationship between brain morphometry and CUD and to further limit type I error, we first conducted zero order correlation analyses to discern which smaller subregions were associated with CUD symptom count. Symptoms are endorsed based on DSM-5 criteria of CUD; therefore, CUD symptom count was selected the outcome to best reflect problematical substance use, rather than CUDIT, which is a screening instrument. ROIs that were significantly correlated with CUD symptom count were retained for further analysis. Next, we conducted a series of multiple linear regressions, with CUD symptom count as the outcome, each lateralized ROI as the predictor, and controlling for alcohol use (i.e., AUDIT; included due to the eligibility criteria and high rates of co-occurrence of alcohol and cannabis use (Harper, Malone, et al., 2021)) and intracranial volume for cortical surface area measures only. False discovery rate (FDR) corrections were used to control type I error inflation for each lateral ROI (Benjamini & Hochberg, 1995). ROIs that survived FDR correction (i.e., q < 0.05) were retained for correlation analyses to assess their functional implications with features of CUD including motives for using cannabis (MMM), depression (PHQ-9), anxiety (GAD-7), trauma (PCL-5), and impulsivity (UPPS-P).

All statistical analyses were conducted in R (version 4.3.1; R Foundation for Statistical Computing, Vienna, Austria).

## Results

### Associations Among OFC Morphometry and CUD Symptoms

Among the seven OFC subregions, lower surface area of the left and right OFC, as well as greater thickness of the left OFC, left pOFC, right 11l, and right 13l were significantly associated a higher number of CUD symptoms (Figure 2). Beyond FDR correction, surface area of the left and right OFC, and cortical thickness of the left OFC, left pOFC, and right 13l continued to be robust in the regressions, controlling for alcohol use and intracranial volume. Measures of cortical surface area of the left and right OFC demonstrated inverse relationships with the number of CUD symptoms, while the remaining measures of thickness were positively associated (Table 2).

**Figure 2.**
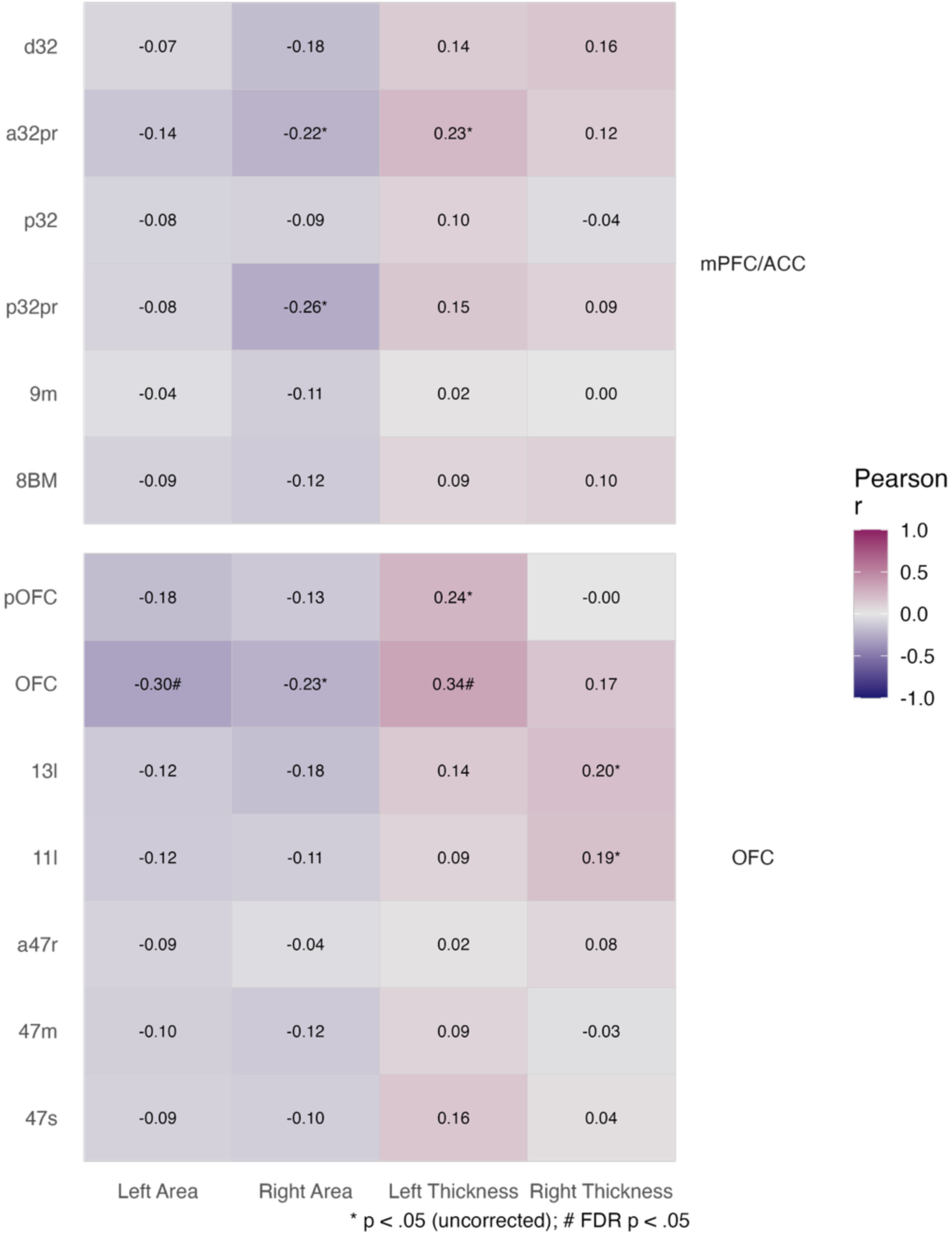
Pearson correlation heat matrix showing associations with orbitofrontal cortex and medial prefrontal/anterior cingulate cortex subregions for surface area and thickness and cannabis use disorder symptom count.

**Table 2.** Associations among orbitofrontal cortex subregions and cannabis use disorder symptom count.

| Region | Hemisphere | Surface Area | P | Thickness | P |
| --- | --- | --- | --- | --- | --- |
| | | $\beta$ (95% CI) | | $\beta$ (95% CI) | |
| OFC | L | **-0.299 (-0.538 to -0.061)** | **0.015\*** | **0.319 (0.135 to 0.503)** | **< 0.001\*** |
|  | R | **-0.274 (-0.531 to -0.017)** | **0.037\*** | 0.167 (-0.019 to 0.353) | 0.078 |
| pOFC | L | -0.127 (-0.368 to 0.113) | 0.296 | **0.262 (0.079 to 0.445)** | **0.005\*** |
|  | R | -0.083 (-0.302 to 0.136) | 0.453 | 0.029 (-0.163 to 0.222) | 0.764 |
| BA 11l | L | 0.032 (-0.211 to 0.275) | 0.795 | 0.081 (-0.108 to 0.270) | 0.396 |
|  | R | 0.072 (-0.191 to 0.334) | 0.590 | 0.179 (-0.008 to 0.365) | 0.060 |
| BA 13l | L | 0.042 (-0.222 to 0.306) | 0.754 | 0.124 (-0.065 to 0.314) | 0.195 |
|  | R | -0.089 (-0.368 to 0.190) | 0.527 | **0.201 (0.016 to 0.386)** | **0.033\*** |
| BA a47r | L | 0.109 (-0.153 to 0.371) | 0.412 | -0.004 (-0.195 to 0.187) | 0.964 |
|  | R | 0.203 (-0.075 to 0.482) | 0.151 | 0.056 (-0.135 to 0.246) | 0.564 |
| BA 47m | L | 0.113 (-0.145 to 0.372) | 0.387 | 0.087 (-0.101 to 0.275) | 0.361 |
|  | R | 0.081 (-0.193 to 0.356) | 0.558 | -0.049 (-0.239 to 0.141) | 0.609 |
| BA 47s | L | 0.093 (-0.166 to 0.351) | 0.479 | 0.152 (-0.035 to 0.338) | 0.110 |
|  | R | 0.042 (-0.219 to 0.302) | 0.753 | 0.036 (-0.152 to 0.225) | 0.704 |

**Notes:** All multiple linear regressions were adjusted for AUDIT. Measures of cortical surface area were adjusted for intracranial volume. Bolded values were included in the FDR correction calculation, with \* designating those retained after false discovery rate correction (i.e., $q < .05$ ). Abbreviations: BA = Brodmann area, OFC = orbitofrontal cortex, pOFC = posterior orbitofrontal cortex.

### Associations Among mPFC/ACC Morphometry and CUD Symptoms

There were only three subregions that were significantly associated with a greater number of CUD symptoms in zero order correlations: lower surface area of the right p32pr and right a32pr, as well as greater thickness of the left a32pr (Figure 2). In the regression analyses, all three regions remained significant beyond FDR correction, demonstrating consistent directionality as the OFC subregions; cortical surface area measures were negatively correlated with the number of CUD symptoms, while cortical thickness was positively correlated (Table 3).

**Table 3.** Associations among medial prefrontal/anterior cingulate cortex subregions and cannabis use disorder symptom count.

| Region | Hemisphere | Surface Area | P | Thickness | P |
| --- | --- | --- | --- | --- | --- |
| | | $\beta$ (95% CI) | | $\beta$ (95% CI) | |
| BA 8BM | L | -0.082 (-0.329 to 0.165) | 0.513 | 0.088 (-0.100 to 0.276) | 0.353 |
|  | R | -0.120 (-0.380 to 0.140) | 0.362 | 0.109 (-0.079 to 0.297) | 0.253 |
| BA 9m | L | -0.038 (-0.313 to 0.236) | 0.782 | 0.013 (-0.175 to 0.202) | 0.888 |
|  | R | -0.101 (-0.384 to 0.181) | 0.478 | 0.009 (-0.180 to 0.198) | 0.922 |
| BA p32pr | L | -0.007 (-0.267 to 0.254) | 0.960 | 0.148 (-0.038 to 0.335) | 0.118 |
|  | R | **-0.285 (-0.532 to -0.037)** | **0.025\*** | 0.108 (-0.081 to 0.296) | 0.259 |
| BA p32 | L | 0.046 (-0.216 to 0.308) | 0.727 | 0.093 (-0.095 to 0.281) | 0.327 |
|  | R | 0.0003 (-0.270 to 0.271) | 0.998 | -0.039 (-0.228 to 0.150) | 0.683 |
| BA a32pr | L | -0.168 (-0.447 to 0.110) | 0.233 | **0.248 (0.064 to 0.431)** | **0.009\*** |
|  | R | **-0.290 (-0.533 to -0.048)** | **0.019\*** | 0.138 (-0.050 to 0.325) | 0.149 |
| BA d32 | L | -0.061 (-0.297 to 0.174) | 0.606 | 0.151 (-0.036 to 0.338) | 0.112 |
|  | R | -0.226 (-0.474 to 0.022) | 0.073 | 0.178 (-0.009 to 0.364) | 0.061 |

**Notes:** All multiple linear regressions were adjusted for AUDIT. Measures of cortical surface area were adjusted for intracranial volume. Bolded values were included in the FDR correction calculation, with \* designating those retained after false discovery rate correction (i.e., $q < .05$ ). Abbreviations: BA = Brodmann area, pr = prime.

### Relationships Between Brain Morphometry and Clinical Characteristics

Among the retained subregions, surface area of the left and right OFC was significantly and negatively correlated with using cannabis to cope (left: r=-0.24, p=0.015; right: r=-0.25, p=0.010). Surface area of the left OFC also was negatively correlated with using cannabis for enhancement (r=-0.21, p=0.036). None of the measures of cortical thickness of OFC subregions were significantly associated with collateral features of CUD.

Among mPFC/ACC subregions, surface area of the right a32pr was negatively correlated with severity of depression (r=-0.24, p=0.011) and trauma (r=-0.24, p=0.012), as well as negative urgency, as measured by the UPPS (r=-0.20, p=0.032). The right p32pr also demonstrated negative correlations, although with the lack of perseverance sub-measure on the UPPS (r=-0.23, p=0.017). Finally, cortical thickness of the left a32pr was positively correlated with severity of depression (r=0.19, p=0.042) and trauma (r=0.19, p=0.049). Figure 3 contains a heat matrix demonstrating associations among OFC and mPFC/ACC subregions.

**Figure 3.**
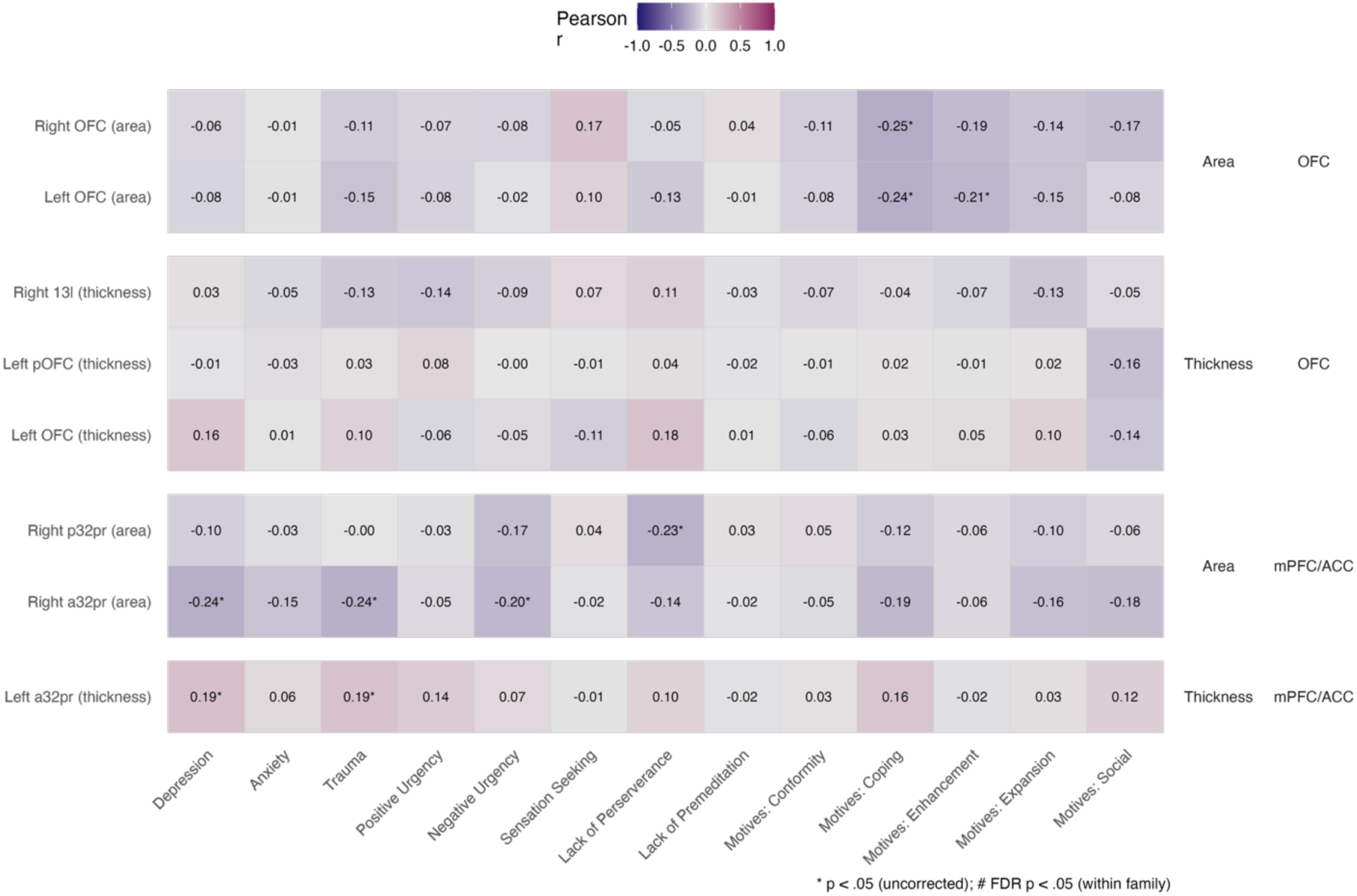
Pearson correlation heat matrices showing associations among orbitofrontal cortex and medial prefrontal/anterior cingulate cortex subregions and collateral cannabis use disorder symptoms for cortical surface area and thickness.

## Discussion

In the present study, we aimed to use the high resolution Glasser atlas to extend the existing literature implicating OFC and mPFC/ACC morphometry correlates of CUD severity in high-risk emerging adults. Among the study participants, high rates of cannabis use were endorsed with 72% of the sample meeting criteria for CUD. Several distinct subregions of both the OFC and mPFC/ACC were identified as associated with CUD severity. Broadly, lower cortical surface area and greater cortical thickness in both regions were associated with a larger number of CUD symptoms. These relationships were further correlated with functional implications of CUD including more severe depression and trauma symptoms, higher levels of impulsivity, and using cannabis to cope or for enhancement. Our findings are consistent with existing literature, while also providing further context and insight into the specific neural mechanisms underlying high-risk cannabis use in emerging adulthood.

Individuals with greater CUD symptom severity demonstrated lower surface area across bilateral OFC subregions, as well as greater cortical thickness in the left OFC, left pOFC, and right 13l. These findings are consistent with previous studies reporting OFC morphometric differences in relation to regular cannabis use and CUD, including evidence of reduced OFC surface area and gray matter volume (Jacobus et al., 2015; Wade et al., 2019). Our finding of greater OFC thickness in relation to CUD severity is consistent with studies reporting thicker lateral OFC among cannabis-using emerging adults and associations between OFC thickness and symptom severity (i.e., Li et al., 2025), highlighting the heterogeneity of structural findings in this region. Functionally, the OFC has a critical role in reward, decision-making, and motivation, processes that are highly relevant to substance use behaviours (Lorenzetti et al., 2019; Harper, Wilson, et al., 2021; Li et al., 2025). Our findings suggest that these functions may be impacted as CUD symptom count increases. Greater CUD severity was also associated with stronger motives to use cannabis both to cope with negative affect and enhance positive affect, patterns that have been observed in prior research (Scarfe et al., 2022; Gex et al., 2024; Halladay et al., 2025; Trinh et al., 2025). The co-occurrence of OFC structural differences and affect-driven cannabis use motives may, therefore, reflect neurobiological disruptions in regions supporting emotion regulation and reward processing, which are particularly salient during emerging adulthood. However, given the cross-sectional design of the study, the presence or magnitude of a causal relationship among these features is unclear.

Greater CUD symptom severity was also associated with structural variations within the mPFC/ACC subregions. Specifically, higher CUD severity was associated with lower surface area in the right p32pr, consistent with prior work reporting reduced mPFC/ACC surface area among cannabis-using emerging adults (Shollenbarger et al., 2015). Conversely, greater CUD severity was associated with increased cortical thickness in the left a32pr, mirroring the pattern observed in OFC subregions. Beyond CUD severity, mPFC/ACC morphology demonstrated robust associations with affective and impulsivity-related features. Lower surface area in right p32pr was associated with greater depressive symptoms, trauma severity, negative urgency, and lack of perseverance, while greater thickness in left a32pr was positively associated with depressive and trauma-related symptoms. Given the central role of mPFC/ACC subregions in affect regulation, cognitive control, and salience processing, these associations are consistent with broader models linking frontal lobe structure to internalizing psychopathology and impulsivity (Maple et al., 2019; Friedman & Robbins, 2022; Sullivan et al., 2022).

The atypical pattern of greater cortical thickness and lower cortical surface area in our findings may be explainable by the microstructure of these two measures. Cortical thickness may increase or decrease based on a number of different reasons including changes in neuronal density, dendritic arborization, neuroplasticity, and reorganization of synapses (Fischl & Dale, 2000; Schüz & Braitenberg, 2001; Aganj et al., 2009; Ledda & Paratcha, 2017), while surface area more directly reflects cortical folding and radial expansion (Panizzon et al., 2009; Winkler et al., 2012). In the current study, our findings may be indicative of two important features of cannabis use in high-risk emerging adults. First, increasing CUD symptom severity in emerging adults may be associated with greater cortical thickness due to changes at the neuronal level. Specifically, greater or more pathological cannabis use may lead to some dendritic morphogenesis to account for reductions or changes in cortical folding—as depicted by reduced cortical surface area. Previous studies have found increased gray matter density among cannabis users, thought to be indicative of denser dendritic branching (Gilman et al., 2014; Brumback et al., 2016). Other studies of cannabis use have reported reduced cortical gyrification in the frontal lobe (Mata et al., 2010; Shollenbarger et al., 2015), suggesting reduced cortical folding and demonstrated as reduced surface area. Overall, our results add to the existing heterogeneity in the literature regarding cortical morphometry in high-risk emerging adults, warranting further investigation and understanding.

### Strengths and Limitations

There are several notable merits of the current study. First, similar studies typically use Desikan or Destrieux atlases for cortical parcellation and segmentation, whereas our current study used the higher resolution Glasser atlas. This provides a much more detailed view of larger regions of the brain, such as the OFC and mPFC/ACC, allowing for greater specificity in brain structure. We are also mindful of several limitations of the present study. First, due to the cross-sectional nature of the current analysis, the regions identified may not be consistent in a longitudinal analysis. As such, we are unable to determine if the observed effects continue beyond the current time point. Alternatively, it is difficult to identify observed effects predate cannabis initiation or are more etiological in nature. Finally, since the Glasser atlas is a surface-based reconstruction, we were unable to assess subcortical regions, such as the nucleus accumbens and caudate. Future studies may benefit from such considerations to better understand the relationship between CUD severity and frontal morphometry in emerging adults.

## Conclusion

Using the Glasser atlas to obtain high-resolution measurements, we found that more severe cannabis use disorder severity was associated with lower cortical surface area and greater thickness across OFC and mPFC/ACC subregions in a sample of high-risk emerging adults. We found further associations with greater internalizing symptoms, impulsivity-related traits, and affect-driven motives for cannabis use, including coping with negative affect and enhancing positive experiences. Our current results offer greater specificity to prior work linking cannabis use to frontal brain structures and highlight dissociable associations between surface area and cortical thickness in regions undergoing continued neurodevelopment during emerging adulthood.

## Data Availability

Data will be made available upon reasonable request and approval by relevant ethical review boards.

## Notes

### Competing Interest Statement

JM serves as a principal and senior scientist at BEAM Diagnostics, Inc., which had no role in the current research or any involvement in any aspect of this work.

### Author Declarations

IRB of the Hamilton Integrated Research Ethics Board at McMaster University gave ethical approval for this work (approval #13763).

### Funding

- Juravinski Research Institute
- Peter Boris Chair in Addictions Research
- Canada Research Chair in Translational Addiction Research: CRC-2020-00170

## References

Aganj, I., Sapiro, G., Parikshak, N., Madsen, S. K., & Thompson, P. M. (2009). Measurement of cortical thickness from MRI by minimum line integrals on soft-classified tissue. Human Brain Mapping, 30(10), 3188–3199. 10.1002/hbm.20740

Albaugh, M. D., Ottino-Gonzalez, J., Sidwell, A., Lepage, C., Juliano, A., Owens, M. M., Chaarani, B., Spechler, P., Fontaine, N., Rioux, P., Lewis, L., Jeon, S., Evans, A., D’Souza, D., Radhakrishnan, R., Banaschewski, T., Bokde, A. L. W., Quinlan, E. B., Conrod, P., … IMAGEN Consortium. (2021). Association of Cannabis Use During Adolescence With Neurodevelopment. JAMA Psychiatry, 78(9), 1031. 10.1001/jamapsychiatry.2021.1258

Babor, T. F., & Robaina, K. (2016). The Alcohol Use Disorders Identification Test (AUDIT): A review of graded severity algorithms and national adaptations. The International Journal of Alcohol and Drug Research, 5(2), 17–24. 10.7895/ijadr.v5i2.222

Benjamini, Y., & Hochberg, Y. (1995). Controlling the false discovery rate: A practical and powerful approach to multiple testing. Journal of the Royal Statistical Society: Series B (Methodological), 57(1), 289–300. 10.1111/j.2517-6161.1995.tb02031.x

Blevins, C. A., Weathers, F. W., Davis, M. T., Witte, T. K., & Domino, J. L. (2015). The Posttraumatic Stress Disorder Checklist for DSM-5 (PCL-5): Development and initial psychometric evaluation. Journal of Traumatic Stress, 28(6), 489–498. 10.1002/jts.22059

Bonn-Miller, M. O., Heinz, A. J., Smith, E. V., Bruno, R., & Adamson, S. (2016). Preliminary Development of a Brief Cannabis Use Disorder Screening Tool: The Cannabis Use Disorder Identification Test Short-Form. Cannabis and Cannabinoid Research, 1(1), 252–261. 10.1089/can.2016.0022

Brumback, T., Castro, N., Jacobus, J., & Tapert, S. (2016). Effects of Marijuana Use on Brain Structure and Function. In International Review of Neurobiology (Vol. 129, pp. 33–65). Elsevier. 10.1016/bs.irn.2016.06.004

Chye, Y., Solowij, N., Suo, C., Batalla, A., Cousijn, J., Goudriaan, A. E., Martin-Santos, R., Whittle, S., Lorenzetti, V., & Yücel, M. (2017). Orbitofrontal and caudate volumes in cannabis users: A multi-site mega-analysis comparing dependent versus non-dependent users. Psychopharmacology, 234(13), 1985–1995. 10.1007/s00213-017-4606-9

Colyer-Patel, K., Romein, C., Kuhns, L., Cousijn, J., & Kroon, E. (2024). Recent Evidence on the Relation Between Cannabis Use, Brain Structure, and Function: Highlights and Challenges. Current Addiction Reports, 11(3), 371–383. 10.1007/s40429-024-00557-z

Cyders, M. A., Littlefield, A. K., Coffey, S., & Karyadi, K. A. (2014). Examination of a short English version of the UPPS-P Impulsive Behavior Scale. Addictive Behaviors, 39(9), 1372–1376. 10.1016/j.addbeh.2014.02.013

Doggett, A., Belisario, K. L., McDonald, A. J., Gohari, M., Leatherdale, S. T., Murphy, J. G., & MacKillop, J. (2025). Evaluating the impact of Canadian cannabis legalization on cannabis use outcomes in emerging adults: Comparisons to a US control sample via a natural experiment. International Journal of Drug Policy, 136, 104686. 10.1016/j.drugpo.2024.104686

Durazzo, T. C., Tosun, D., Buckley, S., Gazdzinski, S., Mon, A., Fryer, S. L., & Meyerhoff, D. J. (2011). Cortical Thickness, Surface Area, and Volume of the Brain Reward System in Alcohol Dependence: Relationships to Relapse and Extended Abstinence: Brain Reward System in Relapse and Abstinence. Alcoholism: Clinical and Experimental Research, 35(6), 1187–1200. 10.1111/j.1530-0277.2011.01452.x

Fischl, B., & Dale, A. M. (2000). Measuring the thickness of the human cerebral cortex from magnetic resonance images. Proceedings of the National Academy of Sciences, 97(20), 11050–11055. 10.1073/pnas.200033797

Friedman, N. P., & Robbins, T. W. (2022). The role of prefrontal cortex in cognitive control and executive function. Neuropsychopharmacology, 47(1), 72–89. 10.1038/s41386-021-01132-0

Gex, K. S., Gückel, T., Wilson, J., Ladd, B. O., & Lee, C. M. (2024). Why People Use Cannabis and Why It Matters: A Narrative Review. Current Addiction Reports, 11(6), 1045–1054. 10.1007/s40429-024-00599-3

Gilman, J. M., Kuster, J. K., Lee, S., Lee, M. J., Kim, B. W., Makris, N., Van Der Kouwe, A., Blood, A. J., & Breiter, H. C. (2014). Cannabis Use Is Quantitatively Associated with Nucleus Accumbens and Amygdala Abnormalities in Young Adult Recreational Users. The Journal of Neuroscience, 34(16), 5529–5538. 10.1523/JNEUROSCI.4745-13.2014

Glasser, M. F., Coalson, T. S., Robinson, E. C., Hacker, C. D., Harwell, J., Yacoub, E., Ugurbil, K., Andersson, J., Beckmann, C. F., Jenkinson, M., Smith, S. M., & Van Essen, D. C. (2016). A multi-modal parcellation of human cerebral cortex. Nature, 536(7615), 171–178. 10.1038/nature18933

González-Roz, A., Castaño, Y., Secades-Villa, R., Janssen, T., Vallejo-Seco, G., & Blanco, C. (2025). Impulsivity traits moderate the longitudinal association between mental health and hazardous cannabis use in emerging adults. Drug and Alcohol Review, 44(4), 1049–1061. 10.1111/dar.14047

Hall, W., Leung, J., & Lynskey, M. (2020). The Effects of Cannabis Use on the Development of Adolescents and Young Adults. Annual Review of Developmental Psychology, 2(1), 461–483. 10.1146/annurev-devpsych-040320-084904

Hall, W., Stjepanović, D., Dawson, D., & Leung, J. (2023). The implementation and public health impacts of cannabis legalization in Canada: A systematic review. Addiction, 118(11), 2062–2072. 10.1111/add.16274

Halladay, J., Belisario, K., McDonald, A., Acuff, S., Doggett, A., Garber, M., Maxwell, A., Murphy, J., & MacKillop, J. (2025). Examining bidirectional associations between cannabis use and internalizing symptoms among high-risk emerging adults: A prospective cohort study. Psychological Medicine, 55, e291. 10.1017/S0033291725101700

Hargreaves, T. L., McIntyre-Wood, C., Vandehei, E., Love, D., Garber, M., Levitt, E. E., Syan, S. K., MacKillop, E., Amlung, M., Sweet, L. H., & MacKillop, J. (2025). Brain structural magnetic resonance imaging predictors of brief intervention response in individuals with alcohol use disorder. Alcohol and Alcoholism (Oxford, Oxfordshire), 60(3), agaf009. 10.1093/alcalc/agaf009

Harper, J., Malone, S. M., Wilson, S., Hunt, R. H., Thomas, K. M., & Iacono, W. G. (2021). The Effects of Alcohol and Cannabis Use on the Cortical Thickness of Cognitive Control and Salience Brain Networks in Emerging Adulthood: A Co-twin Control Study. Biological Psychiatry, 89(10), 1012–1022. 10.1016/j.biopsych.2021.01.006

Harper, J., Wilson, S., Malone, S. M., Hunt, R. H., Thomas, K. M., & Iacono, W. G. (2021). Orbitofrontal cortex thickness and substance use disorders in emerging adulthood: Causal inferences from a co-twin control/discordant twin study. Addiction, 116(9), 2548–2558. 10.1111/add.15447

Harris, P. A., Taylor, R., Thielke, R., Payne, J., Gonzalez, N., & Conde, J. G. (2009). Research electronic data capture (REDCap)—A metadata-driven methodology and workflow process for providing translational research informatics support. Journal of Biomedical Informatics, 42(2), 377–381. 10.1016/j.jbi.2008.08.010

Health Canada. (2023). Canadian Alcohol and Drugs Survey (CADS): Summary of results for 2019. Canadian Alcohol and Drugs Survey (CADS): Summary of Results for 2019. https://www.canada.ca/en/health-canada/services/canadian-alcohol-drugs-survey/2019-summary.html

Jacobus, J., Squeglia, L. M., Meruelo, A. D., Castro, N., Brumback, T., Giedd, J. N., & Tapert, S. F. (2015). Cortical thickness in adolescent marijuana and alcohol users: A three-year prospective study from adolescence to young adulthood. Developmental Cognitive Neuroscience, 16, 101–109. 10.1016/j.dcn.2015.04.006

Jiang, W., Li, G., Liu, H., Shi, F., Wang, T., Shen, C., Shen, H., Lee, S.-W., Hu, D., Wang, W., & Shen, D. (2016). Reduced cortical thickness and increased surface area in antisocial personality disorder. Neuroscience, 337, 143–152. 10.1016/j.neuroscience.2016.08.052

Kroenke, K., Spitzer, R. L., & Williams, J. B. W. (2001). The PHQ-9: Validity of a brief depression severity measure. Journal of General Internal Medicine, 16(9), 606–613. 10.1046/j.1525-1497.2001.016009606.x

Ledda, F., & Paratcha, G. (2017). Mechanisms regulating dendritic arbor patterning. Cellular and Molecular Life Sciences, 74(24), 4511–4537. 10.1007/s00018-017-2588-8

Leung, J., Chan, G. C. K., Hides, L., & Hall, W. D. (2020). What is the prevalence and risk of cannabis use disorders among people who use cannabis? A systematic review and meta-analysis. Addictive Behaviors, 109, 106479. 10.1016/j.addbeh.2020.106479

Li, W., Xu, C., Xu, H., Yin, B., Xu, H., & Li, D. (2025). Abnormal Cortical Thickness Development in Young Adults With Heavy Cannabis Use: A Longitudinal Study. Addiction Biology, 30(5), e70040. 10.1111/adb.70040

Lichenstein, S. D., Manco, N., Cope, L. M., Egbo, L., Garrison, K. A., Hardee, J., Hillmer, A. T., Reeder, K., Stern, E. F., Worhunsky, P., & Yip, S. W. (2022). Systematic review of structural and functional neuroimaging studies of cannabis use in adolescence and emerging adulthood: Evidence from 90 studies and 9441 participants. Neuropsychopharmacology, 47(5), 1000–1028. 10.1038/s41386-021-01226-9

Lorenzetti, V., Chye, Y., Silva, P., Solowij, N., & Roberts, C. A. (2019). Does regular cannabis use affect neuroanatomy? An updated systematic review and meta-analysis of structural neuroimaging studies. European Archives of Psychiatry and Clinical Neuroscience, 269(1), 59–71. 10.1007/s00406-019-00979-1

Maple, K. E., Thomas, A. M., Kangiser, M. M., & Lisdahl, K. M. (2019). Anterior cingulate volume reductions in abstinent adolescent and young adult cannabis users: Association with affective processing deficits. Psychiatry Research: Neuroimaging, 288, 51–59. 10.1016/j.pscychresns.2019.04.011

Mata, I., Perez-Iglesias, R., Roiz-Santiañez, R., Tordesillas-Gutierrez, D., Pazos, A., Gutierrez, A., Vazquez-Barquero, J. L., & Crespo-Facorro, B. (2010). Gyrification brain abnormalities associated with adolescence and early-adulthood cannabis use. Brain Research, 1317, 297–304. 10.1016/j.brainres.2009.12.069

McDonald, A. J., Doggett, A., Belisario, K., Gillard, J., De Jesus, J., Vandehei, E., Lee, L., Halladay, J., & MacKillop, J. (2025). Cannabis Use and Misuse Following Recreational Cannabis Legalization. JAMA Network Open, 8(4), e256551. 10.1001/jamanetworkopen.2025.6551

Nosko, L., Crocker, C. E., & Tibbo, P. G. (2025). Cannabis use in adolescence and young adulthood and its effects on brain structure and function: A scoping review. Frontiers in Psychiatry, 16, 1644105. 10.3389/fpsyt.2025.1644105

Panizzon, M. S., Fennema-Notestine, C., Eyler, L. T., Jernigan, T. L., Prom-Wormley, E., Neale, M., Jacobson, K., Lyons, M. J., Grant, M. D., Franz, C. E., Xian, H., Tsuang, M., Fischl, B., Seidman, L., Dale, A., & Kremen, W. S. (2009). Distinct Genetic Influences on Cortical Surface Area and Cortical Thickness. Cerebral Cortex, 19(11), 2728–2735. 10.1093/cercor/bhp026

Qadeer, R. A., Georgiades, K., Boyle, M. H., & Ferro, M. A. (2019). An Epidemiological Study of Substance Use Disorders Among Emerging and Young Adults. The Canadian Journal of Psychiatry, 64(5), 313–322. 10.1177/0706743718792189

Scarfe, M. L., Muir, C., Rowa, K., Balodis, I., & MacKillop, J. (2022). Getting High or Getting By? An Examination of Cannabis Motives, Cannabis Misuse, and Concurrent Psychopathology in a Sample of General Community Adults. Substance Abuse: Research and Treatment, 16, 11782218221119070. 10.1177/11782218221119070

Schneider, L. H., Pawluk, E. J., Milosevic, I., Shnaider, P., Rowa, K., Antony, M. M., Musielak, N., & McCabe, R. E. (2022). The Diagnostic Assessment Research Tool in action: A preliminary evaluation of a semistructured diagnostic interview for DSM-5 disorders. Psychological Assessment, 34(1), 21–29. 10.1037/pas0001059

Schüz, A., & Braitenberg, V. (2001). Cerebral Cortex: Organization and Function. In International Encyclopedia of the Social & Behavioral Sciences (pp. 1634–1640). Elsevier. 10.1016/B0-08-043076-7/03443-4

Semaan, A., & Khan, M. K. (2026). Neurobiology of Addiction. In StatPearls. StatPearls Publishing. http://www.ncbi.nlm.nih.gov/books/NBK597351/

Shollenbarger, S. G., Price, J., Wieser, J., & Lisdahl, K. (2015). Impact of cannabis use on prefrontal and parietal cortex gyrification and surface area in adolescents and emerging adults. Developmental Cognitive Neuroscience, 16, 46–53. 10.1016/j.dcn.2015.07.004

Simons, J., Correia, C. J., Carey, K. B., & Borsari, B. E. (1998). Validating a five-factor marijuana motives measure: Relations with use, problems, and alcohol motives. Journal of Counseling Psychology, 45(3), 265–273. 10.1037/0022-0167.45.3.265

Spitzer, R. L., Kroenke, K., Williams, J. B. W., & Löwe, B. (2006). A Brief Measure for Assessing Generalized Anxiety Disorder: The GAD-7. Archives of Internal Medicine, 166(10), 1092. 10.1001/archinte.166.10.1092

Sullivan, R. M., Maple, K. E., Wallace, A. L., Thomas, A. M., & Lisdahl, K. M. (2022). Examining Inhibitory Affective Processing Within the Rostral Anterior Cingulate Cortex Among Abstinent Cannabis-Using Adolescents and Young Adults. Frontiers in Psychiatry, 13, 851118. 10.3389/fpsyt.2022.851118

Trinh, C. D., Girard, R., Egan, A., Schick, M. R., & Spillane, N. S. (2025). The Protective Role of Savoring on Coping Motives for Cannabis Use and Consequences. Journal of Psychoactive Drugs, 1–11. 10.1080/02791072.2025.2537043

Wade, N. E., Bagot, K. S., Cota, C. I., Fotros, A., Squeglia, L. M., Meredith, L. R., & Jacobus, J. (2019). Orbitofrontal cortex volume prospectively predicts cannabis and other substance use onset in adolescents. Journal of Psychopharmacology, 33(9), 1124–1131. 10.1177/0269881119855971

Winkler, A. M., Sabuncu, M. R., Yeo, B. T. T., Fischl, B., Greve, D. N., Kochunov, P., Nichols, T. E., Blangero, J., & Glahn, D. C. (2012). Measuring and comparing brain cortical surface area and other areal quantities. NeuroImage, 61(4), 1428–1443. 10.1016/j.neuroimage.2012.03.026

